# *TMEM175, SCARB2* and *CTSB* associations with Parkinson’s disease risk across populations

**DOI:** 10.1101/2025.08.17.25333823

**Authors:** Wenhua Sun, Claudia Schulte, Thomas Gasser, Manuela Tan, the Global Parkinson’s Genetic Program (GP2)

## Abstract

Genome-wide association study of Parkinson’s disease (PD) identified common variants associated with lysosomal mechanism, including *TMEM175, SCARB2*, and *CTSB*. We investigated the association between common and rare variants across populations using cohorts from the Global Parkinson’s Genetics Program (GP2) (33,813 cases and 18,714 controls from 11 ancestries), and the Accelerating Medicines Partnership Parkinson’s Disease (AMP-PD) whole genome sequencing (3,188 cases and 4,104 controls). In the European cohort, we confirmed significant associations with PD risk for all known genetic risk variants across the three genes and *TMEM175 p*.*M393T* as an independent genome-wide significant signal. Additionally, a novel independent signal, *SCARB2* rs11547135, was detected. The burden analysis linked PD to *SCARB2* in African American, Ashkenazi Jewish and East Asian cohorts. Single variants-based tests identified rare missense and synonymous variants in *SCARB2* in several populations. Our study reinforces the association of lysosomal genetic variants with PD risk, revealing genetic heterogeneity across populations.

## Introduction

Glucocerebrosidase (GCase), encoded by *GBA1*, is a lysosomal enzyme critical for maintaining lysosomal protein and lipid homeostasis. Gcase deficiency causes accumulation of alpha-synuclein(α-syn)^1^ and variants in *GBA1* are recognized as a common genetic risk factor for PD^2^. The latest genome-wide association study (GWAS) in European derived populations identified common genetic variants associated with lysosomal dysfunction that contribute to Parkinson’s disease (PD) risk, with *GBA1* (2019 PD GWAS, rs76763715 (c.1226A>G; p.Asn409Ser, odds ratio [OR] = 2.11) emerging as a key locus^3,4^. Genetic variants in other lysosome-related genes (including transmembrane protein 175 [*TMEM175*], Scavenger Receptor Class B Member 2 [*SCARB2*] and Cathepsin B [*CTSB*]) are also associated with PD risk and age at onset^5-7^. These candidate genes were prioritized in this study based on (1) *GBA1*’s role as a major lysosomal driver of PD pathogenesis; (2) functional convergence of *TMEM175, SCARB2*, and *CTSB* with GCase activity and lysosomal function —either through direct interaction (*SCARB2*)^8^, pH regulation (*TMEM175*)^9,10^, or cathepsin B(*CTSB*) in mediating prosaposin cleavage to form saposin C, the lysosomal coactivator of GCase^11,12^. In addition to their mechanistic relevance, these three genes were identified as genome-wide significant loci in large-scale GWAS of PD^3^, further supporting their contribution to disease risk. Together, these genes interact to provide a cohesive framework to dissect lysosomal mechanisms in PD. Their distinct roles are detailed below.

The *TMEM175* gene encodes a proton-selective ion channel located on lysosomal membranes. It mediates the lysosomal H+ leak that balances vacuolar-type H+-ATPase (V-ATPase) activity which helps maintain lysosomal pH homeostasis^10^. Two common coding variants in the *TMEM175* gene, rs34311866 (p.M393T) and rs34884217 (p.Q65P), show opposite effects on PD susceptibility in several populations^6,13,14^. The GWAS identified p.M393T associated with increased risk (2019 PD GWAS, rs34311866, OR =1.237)^3^ and earlier age of onset of PD^13,15^.

*SCARB2* is an intracellular receptor which shuffles GCase from the endoplasmic reticulum to the lysosome. Variants in this gene may lead to functional and structural lysosomal dysfunction^8^. Two common intronic variants, rs6812193 and rs6825004 (2019 PD GWAS, rs6825004, OR = 0.9397; rs6812193, OR = 1.09), have been associated with the risk of PD in several genetic studies^3,8,16-18^.

*CTSB* is a cysteine protease, which plays an essential role in lysosomal degradation of α-syn^11^. The *CTSB* locus harbors an intronic variant (rs1293298) which is a common genetic risk factor for PD (2019 PD GWAS, OR = 0.9112)^3^. Interestingly, this variant also modifies PD risk in *GBA1* carriers, suggesting an interaction between *GBA1* and *CTSB*^5^. This variant lowers the penetrance of *GBA1* mutations, reducing the risk of PD in carriers.

To overcome limitations of prior studies—including insufficient sample sizes for robust statistical inference, ancestry heterogeneity or Eurocentric cohort bias, and fragmented assessments of lysosomal genes contributions, we leveraged large-scale cohorts of genotyping and whole genome sequencing (WGS) data from AMP-PD (The Accelerating Medicines Partnership Parkinson’s disease) and GP2 (Global Parkinson’s Genetics Program) and investigated association of common and rare variants in lysosomal related genes (*TMEM175, SCARB2, CTSB* and *GBA1*) with PD across 11 populations.

## Methods

### 1. Subjects

To validate the association between common variants in lysosomal genes from the reported GWA study of PD^3^, large-scale Neurobooster genotyping imputed data obtained from GP2 release 10 (https://gp2.org/the-components-of-gp2s-8th-data-release/) was analysed, which contains 33,813 cases and 18,714 controls in 11 ancestries: GP2-African American (GP2-AAC: 369 cases, 756 controls), GP2-African (GP2-AFR: 1,147 cases, 2,179 controls), GP2-Ashkenazi Jewish (GP2-AJ: 1,285 cases, 408 controls), GP2-Latino and indigenous Americas (GP2-AMR: 1,917 cases, 1,402 controls), GP2-Central Asian (GP2-CAS: 734 cases, 578 controls), GP2-East Asian (GP2-EAS: 2,377 cases, 2,598 controls), GP2-European (GP2-EUR: 24,208 cases, 9,662 controls), GP2-Finnish (GP2-FIN: 80 cases, 11 controls) population isolate; GP2-Middle Eastern (GP2-MDE: 724 cases, 551 controls), GP2-South Asian (GP2-SAS: 309 cases, 261 controls), GP2-Complex Admixture History (GP2-CAH: 663 cases, 318 controls). To minimize potential bias, individuals carrying *GBA1* risk variants were excluded from all ancestries. Quality control analyses at a sample and variant level for this GP2 dataset are described elsewhere (https://github.com/GP2code/GenoTools)^19^. To validate the association between rare variants in *GBA1* and PD risk, WGS data from AMP-PD release 4 (https://www.amp-pd.org/news/gp2-release-notes-november-2023) was analysed, which contains 3,188 cases and 4,104 controls from EUR ancestry. Information about the quality control steps performed on the AMP-PD dataset can be found at: https://amp-pd.org/whole-genome-data. Cases and controls with missing covariates (including gender and age) were removed.

### 2. Gene region extraction and variants annotation

Regions of each gene were extracted with PLINK 2.0 based on the gene locations according to the Ensembl genome browser (https://www.ensembl.org/Homo_sapiens/), including 50 kb downstream and upstream of the genes. Variants were annotated using ANNOVAR^20^, and variants were defined as (1) exonic, (2) non-synonymous and loss of function (Nonsyn and LoF). LoF included stopgain, stoploss and frameshift variants.

### 3. Statistical analysis

Imputed genotype data often struggles to accurately capture rare variants (MAF <1%) in complex genomic regions such as *GBA1* locus because of poor imputation quality and genomic complexity. Therefore, GP2 Neurobooster data was used to assess both common variants and rare variants in the genes *TMEM175, SCARB2* and *CTSB*, while AMP-PD WGS data was used to conduct rare variant association analysis in the *GBA1* gene.

Association analyses were performed with PLINK 2.0^21^, adjusted by gender, age and the first five genetic principal components (PCs) in each ancestry.

Conditional and joint (COJO) analysis was conducted to determine if there are independent signals per ancestry with genome-wide complex trait analysis (GCTA) software version 1.94.1^22,23^. To identify gene-wide significant signals in the absence of genome-wide significance (p <5×10^⁻8^), we established gene-specific significant thresholds through linkage disequilibrium (LD) pruning followed by Bonferroni correction. LD pruning was performed to define a p-value threshold for the COJO analysis in GP2 NeuroBooster array (parameters: --indep-pairwise 50 50 0.5). We identified 107 independent variants in *TMEM175*, 141 variants in *SCARB2* and 116 variants in *CTSB*. Therefore, the thresholds for p values which were used in conditional analysis were: 4.67e-4 (0.05/107, *TMEM175*), 3.55e-4 (0.05/141, *SCARB2*), 4.31e-4 (0.05/116, *CTSB*). In addition, LDpair tool (https://ldlink.nih.gov/?tab=ldpair) was used to validate if these signals were independent of known GWAS hits. Regional plots were generated with the online tool LocusZoom.js (https://statgen.github.io/localzoom/)^24^.

WGS data obtained from AMP-PD release 4.0 (https://www.amp-pd.org/news/gp2-release-notes-november-2023) and GP2 NeuroBooster array dataset was used separately to assess whether *TMEM175, SCARB2, CTSB* and *GBA1* have an excessive genetic burden on PD. Gene-based burden tests were performed using RVTESTS^25^ to assess the cumulative effects of rare variants on the risk for PD. Analyses were performed twice for each gene, once for all exonic variants and once for the stricter subset of Nonsyn+LoF variants. Benjamini–Hochberg procedure was used to adjust the raw P-values for multiple testing. Furthermore, single-variant-based tests were conducted with RVTESTS to find single statistically significant variants which have an impact on PD risk. All burden analyses were adjusted for age, gender and PC1-PC5.

All analyses were performed using R statistical language, Python and linux programming language on Terra (https://terra.bio/) and Verily workbench (https://workbench.verily.com/).

Our overall analytic approach is illustrated in Figure 1.

**Figure 1.**
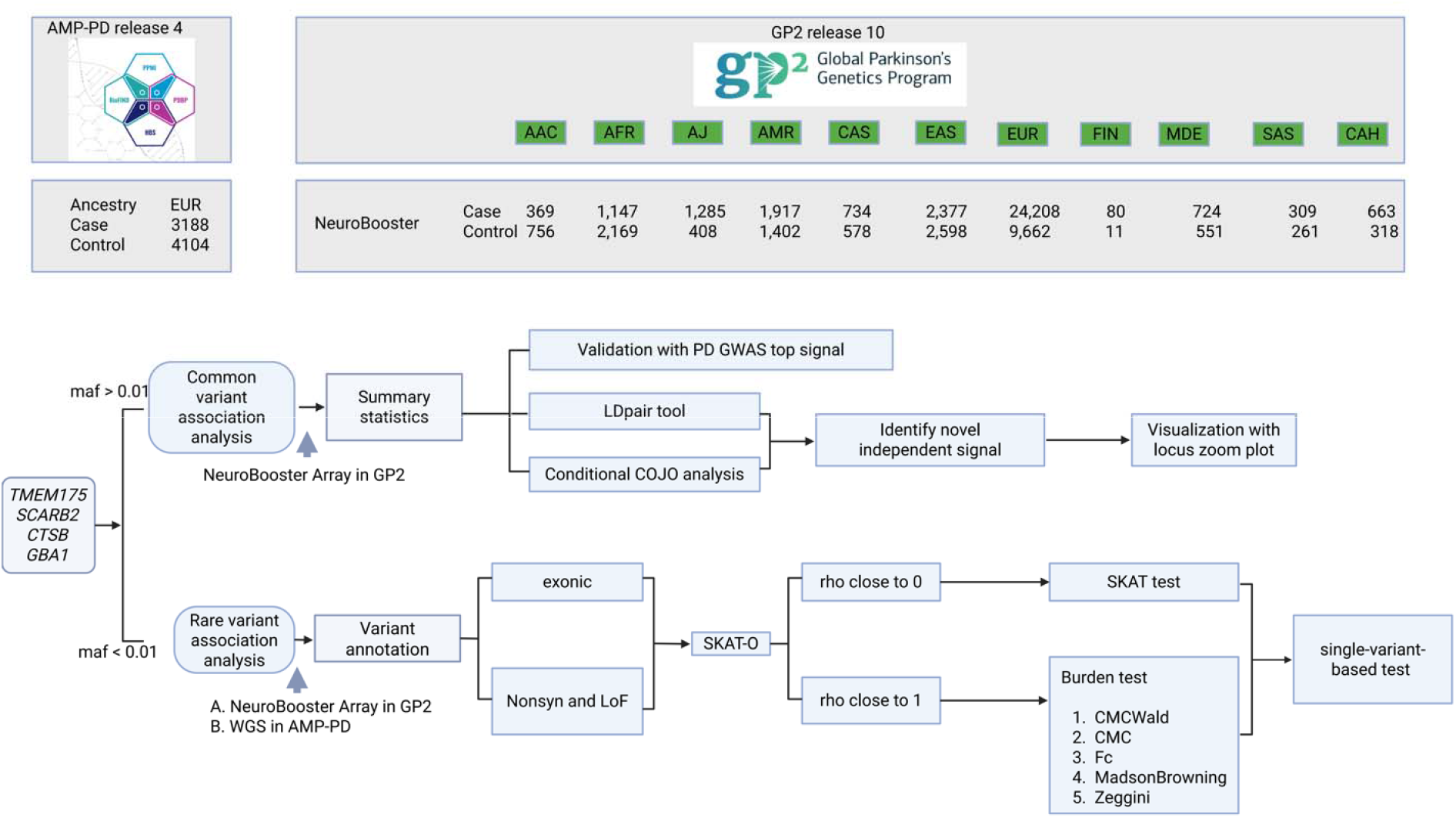
General overview of analysis. Abbreviations, maf, minor allele frequency; WGS, whole genome sequencing; Nonsyn and LoF, non-synonymous variants.

## Results

### 1. Association analysis of known common risk variants in TMEM175, SCARB2, and CTSB across 11 populations

#### (1) GP2-EUR cohort

To validate previously reported GWAS hits, we tested the association of the following variants with PD risk in the GP2-EUR cohort: *TMEM175* [p.M393T(rs34311866) and p.Q65P(rs34884217)], *SCARB2* (rs6812193 and rs6825004), and *CTSB* (rs1293298). We replicated previous findings that both *TMEM175* variants, p.M393T (rs34311866) reached genome-wide significance and p.Q65P (rs34884217) was nominally significant in the GP2-EUR cohort [p value = 6.07e-16, OR (95%CI) = 1.25(1.19-1.32) and p value = 1.70e-4, OR (95%CI) = 0.88 (0.82-0.94).]

Both variants in *SCARB2*, rs6812193[p value =7.81e-03, OR (95%CI) = 0.94(0.90-0.98)] and rs6825004[p value = 0.036, OR (95%CI) = 0.95(0.91-1.00)], showed nominal significance in the GP2-EUR cohort, though neither reached genome-wide significance (Supplementary Table 1).

**Table 1.** Association analysis of *TMEM175* p.M393T and p.Q65P across all 11 populations

| Ancestry | *TMEM175* p.M393T<br>(rs34311866, 4:958159: T:C) |  |  |  | *TMEM175* p.Q65P<br>(rs34884217, 4:950422: A:C) |  |  |  |
| --- | --- | --- | --- | --- | --- | --- | --- | --- |
|  | MAF (cases) | MAF (controls) | P value | OR (95%CI) | MAF (cases) | MAF (controls) | P value | OR (95%CI) |
| AAC | 0.051 | 0.051 | 0.18 | 0.71 (0.43-1.17) | 0.027 | 0.024 | 0.75 | 0.90 (0.45-1.77) |
| AFR | NA | NA | NA | NA | NA | NA | NA | NA |
| AJ | 0.35 | 0.28 | 2.51e-03* | 1.43 (1.13-1.80) | 0.082 | 0.10 | 0.042* | 0.70 (0.49-0.99) |
| AMR | 0.088 | 0.040 | 0.020* | 1.36 (1.05-1.75) | 0.062 | 0.032 | 0.85 | 1.03 (0.77-1.36) |
| CAS | 0.22 | 0.22 | 0.64 | 1.06 (0.82-1.37) | 0.037 | 0.029 | 0.46 | 1.22 (0.72-2.05) |
| EAS | 0.15 | 0.14 | 0.086 | 1.14 (0.98-1.34) | NA | NA | NA | NA |
| EUR | 0.21 | 0.18 | 6.07e-16* | 1.25(1.19-1.32) | 0.093 | 0.11 | 1.70e-04* | 0.88(0.82-0.94) |
| FIN | 0.18 | 0.27 | 0.41 | 0.29(0.015-5.56) | 0.075 | 0 | 0.57 | 0.51(0.052-5.05) |
| MDE | 0.23 | 0.24 | 0.019* | 1.30 (1.04-1.63) | 0.061 | 0.087 | 0.019* | 0.66 (0.47-0.93) |
| SAS | 0.38 | 0.28 | 8.88e-04* | 1.83(1.28-2.61) | 0.024 | 0.019 | 0.41 | 1.75(0.46-6.57) |
| CAH | 0.15 | 0.16 | 0.44 | 0.88(0.64-1.22) | 0.070 | 0.050 | 0.37 | 1.24 (0.77-1.98) |

Abbreviations: AAC, African American; AFR, African; AJ, Ashkenazi Jewish; AMR, Latino and indigenous Americas; CAS, Central Asian; EAS, East Asian; EUR, European; FIN, Finnish population isolate; MDE, Middle Eastern; SAS, South Asian; CAH, Complex Admixture History. MAF, minor allele frequency. OR, odds ratio. CI, Confidence Interval. NA not assessed because MAF < 0.01. \* p value < 0.05.
Location of variants are based on GRCh38/hg38.

An association between *CTSB* rs1293298 and PD was found to be nominally significant in the GP2-EUR cohort [p value = 5.82e-04, OR (95%CI) = 0.92(0.87-0.96)] (Supplementary Table 2).

**Table 2.**
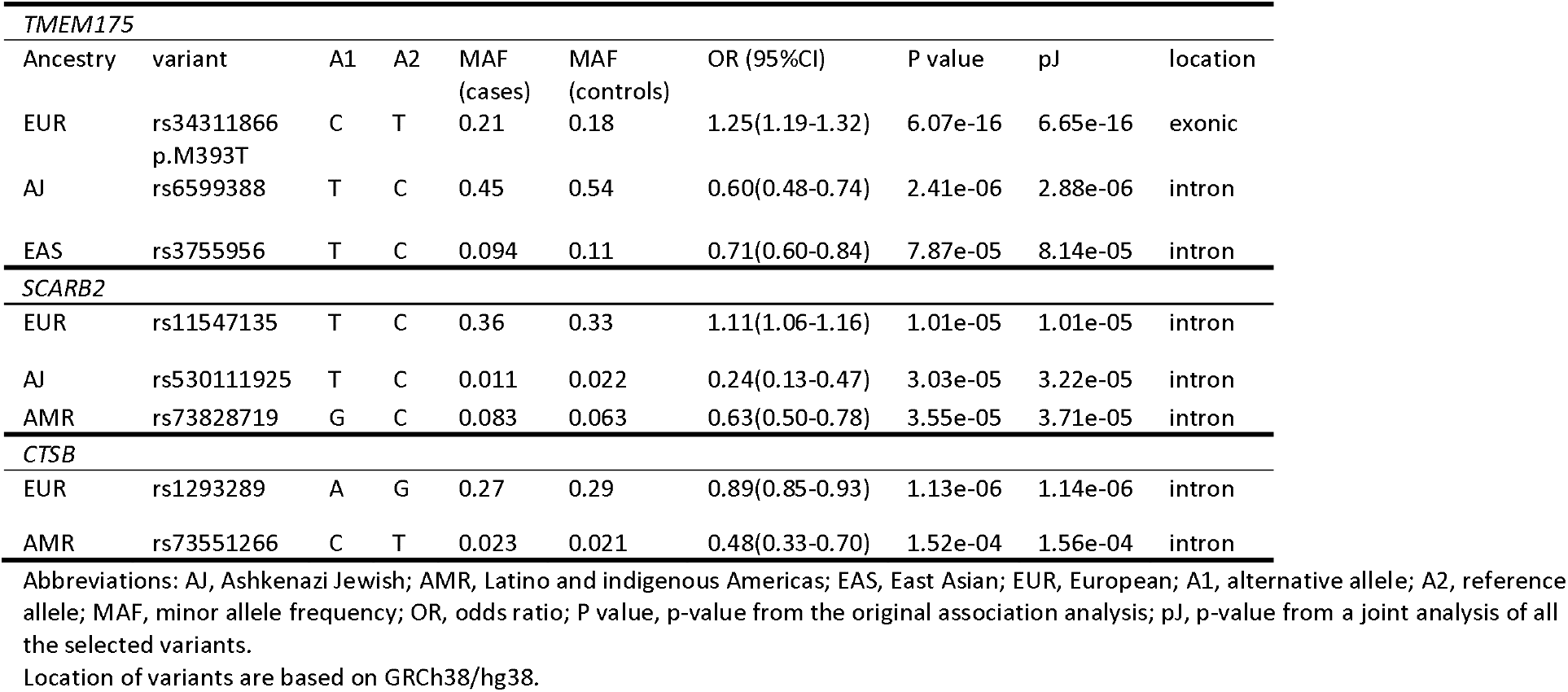
Independent signals in *TMEM175, SCARB2* and *CTSB*

| *TMEM175* |  |  |  |  |  |  |  |  |  |
| --- | --- | --- | --- | --- | --- | --- | --- | --- | --- |
| Ancestry | variant | A1 | A2 | MAF<br>(cases) | MAF<br>(controls) | OR (95%CI) | P value | pJ | location |
| EUR | rs34311866<br>p.M393T | C | T | 0.21 | 0.18 | 1.25(1.19-1.32) | 6.07e-16 | 6.65e-16 | exonic |
| AJ | rs6599388 | T | C | 0.45 | 0.54 | 0.60(0.48-0.74) | 2.41e-06 | 2.88e-06 | intron |
| EAS | rs3755956 | T | C | 0.094 | 0.11 | 0.71(0.60-0.84) | 7.87e-05 | 8.14e-05 | intron |
| *SCARB2* |  |  |  |  |  |  |  |  |  |
| EUR | rs11547135 | T | C | 0.36 | 0.33 | 1.11(1.06-1.16) | 1.01e-05 | 1.01e-05 | intron |
| AJ | rs530111925 | T | C | 0.011 | 0.022 | 0.24(0.13-0.47) | 3.03e-05 | 3.22e-05 | intron |
| AMR | rs73828719 | G | C | 0.083 | 0.063 | 0.63(0.50-0.78) | 3.55e-05 | 3.71e-05 | intron |
| *CTSB* |  |  |  |  |  |  |  |  |  |
| EUR | rs1293289 | A | G | 0.27 | 0.29 | 0.89(0.85-0.93) | 1.13e-06 | 1.14e-06 | intron |
| AMR | rs73551266 | C | T | 0.023 | 0.021 | 0.48(0.33-0.70) | 1.52e-04 | 1.56e-04 | intron |

Abbreviations: AJ, Ashkenazi Jewish; AMR, Latino and indigenous Americas; EAS, East Asian; EUR, European; A1, alternative allele; A2, reference allele; MAF, minor allele frequency; OR, odds ratio; P value, p-value from the original association analysis; pJ, p-value from a joint analysis of all the selected variants.
Location of variants are based on GRCh38/hg38.

#### (2) Other GP2 cohorts

The known risk factor in *TMEM175* p.M393T (rs34311866) was found to be significantly associated with PD in the GP2-SAS cohort [p value = 8.88e-04, OR (95%CI) = 1.83(1.28-2.61)], GP2-MDE cohort [p value = 0.019, OR (95%CI) = 1.30(1.04-1.63)], and GP2-AJ cohort [p value =2.51e-03, OR (95%CI) = 1.43 (1.13-1.80)]. In addition, *TMEM175* p.Q65P (rs34884217) was significant in the GP2-MDE cohort [p value = 0.019, OR (95%CI) = 0.66(0.47-0.93))] and GP2-AJ cohort [p value = 0.042, OR (95%CI) = 0.70(0.49-0.99))]. Notably, the *TMEM175* p.M393T had a minor allele frequency (MAF) below 0.01 in the African ancestry and p.Q65P variant in the African and East Asian ancestry and were therefore not analysed (Table 1). In addition, the *SCARB2* rs6825004 variant was significantly associated with reduced PD risk in the GP2-CAH cohort [p value = 0.042, OR (95%CI) = 0.78(0.62-0.99)] and in the GP2-MDE cohort [p value = 0.023, OR (95%CI) = 0.80(0.66-0.97)] (Supplementary Table 1). A protective association between *SCARB2* rs6812193 and PD risk was observed in the GP2-AMR cohort [p value = 0.020, OR (95%CI) = 0.83(0.71-0.97)]. *CTSB* rs1293298 were significantly associated with PD risk in GP2-CAH cohort [p value = 0.021, OR (95%CI) = 0.73(0.56-0.95)] (Supplementary Table 2).

### 2. Identification of independent signals with conditional analysis across 11 populations

#### (1)TMEM175

The GWAS signal in the *TMEM175* gene, p.M393T (rs34311866) was validated as an independent signal in the GP2-EUR cohort (pJ = 6.65e-16) (Table2; Supplementary Figure 1). In the GP2-AJ cohort, rs6599388 (pJ = 2.88e-06) was identified as an independent signal (Table 2; Supplementary Figure 2). We found rs6599388 was correlated with the previously reported variant rs34311866 (D’ = 0.87, R^2^ = 0.39, p <0.0001) in the GP2-AJ cohort. In the GP2-EAS cohort, another signal rs3755956 was identified as a novel independent signal, showing no correlation with rs34311866(D’ = 0.12, R^2^ = 0.0003, p = 0.59) (Table 2; Supplementary Figure 3).

#### (2) SCARB2

We identified one independent signal, rs11547135(pJ = 1.01e-05), in *SCARB2* in the GP2-EUR cohort (Table 2; Supplementary Figure 4). rs11547135 was not correlated with previously reported variants rs6812193(D’ = 0.43, R^2^ = 0.056, p <0.0001) and rs6825004(D’ = 0.27, R^2^ = 0.073, p <0.0001). One additional novel independent genetic variants in *SCARB2*, rs530111925(pJ = 3.22e-05) was identified in the GP2-AJ cohort(Table2; Supplementary Figure 5) and this variant was not correlated with rs6812193(D’ = 0.31,R^2^ = 0.0027, p <0.0001) and rs6825004(D’ = 0.14, R^2^ = 0.00035, p = 0.12). In the GP2-AMR cohort, one independent signal, rs73828719(pJ = 3.71e-05) was correlated with rs6812193(D’ = 0.87,R^2^ = 0.26, p <0.0001) but not correlated with rs6825004 (D’ = 0.19, R^2^ = 0.0028, p <0.0001). (Table 2; Supplementary Figure 6).

#### (3) CTSB

We identified one independent signal in *CTSB*, rs1293289(pJ = 1.14e-06), in GP2-EUR cohort, but rs1293289 was correlated with the known PD GWAS hit rs1293298 (D’ = 0.97, R^2^ = 0.80, p <0.0001) (Table 2; Supplementary Figure 7). In the GP2-AMR cohort, one independent novel genetic variants in *CTSB*, rs73551266(pJ = 1.56e-04) was identified (Table 2; Supplementary Figure 8) and rs73551266 was independent of rs1293298(D’ = 0.0038, R^2^ = 0.00, p = 0.96).

### 3. Burden analysis with GP2 Neurobooster array and AMP-PD datasets

To test if an aggregate burden of rare variants in *TMEM175, SCARB2* and *CTSB* contributes to the risk of PD, SKAT-O analysis was conducted within the GP2 Neurobooster array data across 11 populations (Table 3).

**Table 3.** Skat-O analysis of *TMEM175, SCARB2* and *CTSB*

| Ancestry | *TMEM175* |  |  |  | *SCARB2* |  |  | *CTSB* |  |  |
| --- | --- | --- | --- | --- | --- | --- | --- | --- | --- | --- |
|  | Variants | Num<br>Var | rho | P<br>value | NumVar | rho | P<br>value | NumVar | rho | P<br>value |
| AAC | exonic | 59 | 0 | 0.63 | 21 | 0 | 1.90e-03* | 28 | 0 | 0.75 |
|  | Nonsyn and LoF | 38 | 0 | 0.63 | 12 | 0 | 0.050 | 15 | 0 | 0.55 |
| AFR | exonic | 76 | 1 | 0.74 | 19 | 1 | 0.78 | 46 | 0 | 0.83 |
|  | Nonsyn and LoF | 47 | 1 | 0.74 | 10 | 1 | 0.55 | 25 | 0.30 | 0.29 |
| AJ | exonic | 24 | 0 | 0.75 | 5 | 0 | 0.021* | 13 | 1 | 0.88 |
|  | Nonsyn and LoF | 15 | 1 | 0.75 | 4 | NA | NA | 11 | 1 | 0.88 |
| AMR | exonic | 72 | 0 | 0.83 | 18 | 0 | 0.85 | 39 | 0 | 0.75 |
|  | Nonsyn and LoF | 42 | 1 | 0.83 | 14 | 0 | 0.85 | 27 | 0 | 0.75 |
| CAS | exonic | 39 | 1 | 0.26 | 15 | 0 | 0.13 | 18 | 1 | 0.86 |
|  | Nonsyn and LoF | 26 | 0 | 0.26 | 10 | 0.2 | 0.091 | 12 | 1 | 0.86 |
| EAS | exonic | 52 | 0 | 0.86 | 17 | 0 | 0.024* | 24 | 1 | 1 |
|  | Nonsyn and LoF | 34 | 0 | 0.80 | 12 | 0 | 0.024* | 17 | 1 | 1 |
| EUR | exonic | 128 | 0 | 0.80 | 43 | 0 | 0.54 | 93 | 0 | 0.57 |
|  | Nonsyn and LoF | 69 | 1 | 0.86 | 27 | 0 | 0.54 | 59 | 1 | 0.57 |
| FIN | exonic | 4 | NA | NA | NA | NA | NA | NA | NA | NA |
|  | Nonsyn and LoF | 2 | NA | NA | NA | NA | NA | NA | NA | NA |
| MDE | exonic | 47 | 0 | 0.42 | 21 | 1 | 1 | 24 | 1 | 0.57 |
|  | Nonsyn and LoF | 24 | 0 | 0.13 | 15 | 1 | 1 | 15 | 1 | 0.57 |
| SAS | exonic | 25 | 0 | 0.55 | 10 | 0 | 0.29 | 19 | 1 | 0.64 |
|  | Nonsyn and LoF | 18 | 1 | 0.55 | 7 | 0.8 | 0.29 | 12 | 0 | 0.64 |
| CAH | exonic | 47 | 1 | 0.64 | 16 | 1 | 1 | 28 | 0 | 0.82 |
|  | Nonsyn and LoF | 30 | 0 | 0.64 | 9 | 1 | 1 | 14 | 1 | 0.82 |

Abbreviations: AAC, African American; AFR, African; AJ, Ashkenazi Jewish; AMR, Latino and indigenous Americas; CAS, Central Asian; EAS, East Asian; EUR, European; FIN, Finnish population isolate; MDE, Middle Eastern; SAS, South Asian; CAH, Complex Admixture History; Nonsyn, non-synonymous variants; LoF, Loss of function; NA, not available. NumVar, number of variants.

#### (1) TMEM175 and CTSB

No significant associations were detected for rare exonic and Nonsyn and LoF variants in *TMEM175* and *CTSB* across 11 GP2 cohorts.

#### (2) SCARB2

Similarly, a SKAT-O analysis for *SCARB2* was run to detect a potential genetic burden in PD cases versus controls (Table 3). There was no significant association in the GP2-EUR cohort either for exonic (N variants = 43, rho=0, p = 0.54) or for Nonsyn and LoF variants (N variants = 27, rho =0, p = 0.54). However, significant associations were detected for exonic variants in the GP2-AAC cohort (N variants = 21, rho=0, p = 1.90e-03) and in the GP2-AJ cohort (N variants = 5, rho=0, p = 0.021). In the GP2-EAS cohort, there was a significant genetic burden in PD for the exonic and Nonsyn and LoF subsets (exonic: N variants = 17, rho=0, p = 0.024; Nonsyn and LoF: N variants = 12, rho=0, p = 0.024) (Table 3).

Subsequently, a SKAT test and different burden tests were performed to further validate SKAT-O results according to the rho value (Supplementary table 3). While a rho value close to 1 corresponds to a pure burden (collapsing) test where all variants have concordant directions of effect, a rho value close to 0 corresponds to a pure SKAT (kernel-based) test^26^.

Hence, a SKAT test was conducted next, and significant associations were detected in the exonic subset of the GP2-AAC cohort (p = 8.90e-04) and GP2-AJ cohort (p = 0.021) (Supplementary table 3). In the GP2-EAS cohort, the SKAT test showed an association signal (exonic: p = 0.013; Nonsyn and LoF: 0.011) (Supplementary table 3).

To determine which single variants may be driving the observed association with PD risk, a single-variant-based test was conducted (Table 4). Five novel variants related with increased risk of PD (rs149997095, effect allele=G, effect = 17.37, SE = 5.11, p = 6.85e-04; rs957776724, effect allele=C, effect = 48.17, SE = 8.86, p = 5.38e-08; rs143518519, effect allele=A, effect = 16.54, SE = 5.11, p = 1.23e-03; rs147159813, effect allele=T, effect = 11.52, SE = 4.43, p = 9.29e-03; rs35069772, effect allele=A, effect = 42.50, SE = 8.85, p = 1.58e-06) in the GP2-AAC cohort were identified(Table 4). One novel missense variant (rs143655258, effect allele=C, effect = -5.25, SE = 2.27, p = 0.021) had a protective impact on PD risk in the GP2-AJ cohort(Table 4). A novel variant, rs773017713, was found to decrease the risk of PD (effect allele=T, effect = -4.31, SE = 1.78, p = 0.015) in the GP2-EAS cohort (Table 4) and another variant, rs117600063, increased PD risk (effect allele=A, effect = 2.05, SE = 0.91, p = 0.024) in the GP2-EAS cohort (Table 4).

**Table 4.**
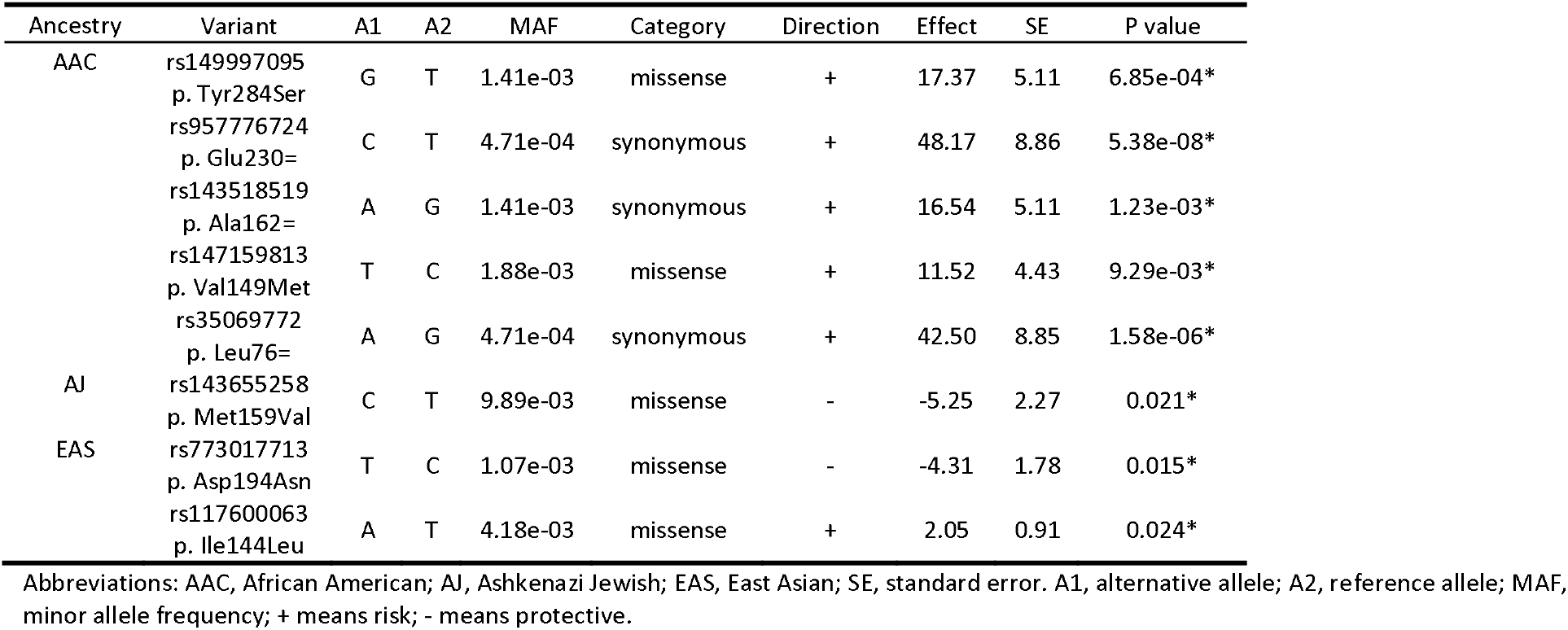
Significant signals with single variant burden analysis (*SCARB2*)

#### (3) GBA1

The AMP-PD WGS dataset was leveraged to undertake a burden analysis of *GBA1*. There was suggestive evidence for an increased burden of *GBA1* rare variants associated with PD in the European population (exonic, rho =1, p = 8.60e-03; Nonsyn and LoF, rho = 1, p = 0.020) (Supplementary table 4A). One known variant, rs76763715 (*GBA1* p. Asn409Ser), was shown to be associated with PD risk (effect allele=T, effect = 1.11, SE = 0.39, p = 4.00e-03) (Supplementary table 4C).

## Discussion

We show that previous GWAS hits of three lysosomal related genes (*TMEM175, SCARB2* and *CTSB*) were successfully replicated using large scale genotyping array data. All previously detected variants were significant in the cohort of European ancestry and in some additional ancestry cohorts. The GCTA-COJO analysis was performed across all 11 cohorts to identify novel independent signals in these three lysosome-related genes. Significant associations were observed in GP2-EUR, GP2-AJ, GP2-EAS, and GP2-AMR cohorts. Furthermore, SKAT-O analysis revealed genetic burdens for lysosomal related genes (e.g. *GBA1* and *SCARB2*), with each gene showing significant associations in distinct cohorts. Collapsed burden tests and SKAT tests were further used to validate these findings. Moreover, we detected single rare variants contributing to PD risk through single-variant-based tests.

### TMEM175

The rs34311866 missense variant p.M393T in the *TMEM175* gene was the most strongly associated signal in the region. The association reached genome-wide significance and *TMEM175* rs34311866 was identified as a single independent risk factor of PD in the European population, aligning with previous reports^3^. In addition, it was associated with PD in Ashkenazi Jewish, Latino and indigenous Americas, Middle Eastern and South Asian populations, which could be possibly due to a haplotype block including rs34311866 variant in these other populations. Another common variant rs34884217 p.Q65P, as a secondary signal in the same locus, was found to be associated with reduced PD risk in a previous study^27^. In this study, the frequency of the protective allele was also significantly higher in the controls than in cases across Ashkenazi Jewish, European and Middle Eastern populations. Notably, due to the minor allele frequency (MAF) threshold of 0.01 for common variants, *TMEM175* rs34311866 and rs34884217 in the African, and *TMEM175* rs34884217 in the East Asian populations, were not included in the analysis in these populations.

We went further to identify independent signals in *TMEM175* using GCTA-COJO analysis in other populations. Genotypes at rs6599388 in the *TMEM175* gene was detected as an independent intronic variant for PD risk with GCTA-COJO in the Ashkenazi Jewish population. Of note, *TMEM175* rs6599388 was correlated with rs34311866. It was also previously reported to be associated with PD in GWA studies in the European population (2011 PD GWAS, rs6599388, odds ratio [OR] = 1.16)^28^. In the East Asian population, conditional analysis identified rs3755956 as a novel, independent association signal at the *TMEM175* locus.

No statistically significant associations of rare variants in the *TMEM175* gene were found for any ancestral populations. A previous study did not show significant results in 2,657 patients and 3,647 controls in a *TMEM175* burden analysis^29^. Our finding was consistent with this previous study, supported by sufficient statistical power, suggesting that a cumulative burden of rare variants in the *TMEM175* gene does not contribute significantly to PD risk.

### SCARB2

Regarding *SCARB2*, our findings support previous reports that genotypes at rs6812193 and rs6825004 in the *SCARB2* gene were found associated with PD in the European ancestry^3,8,17,18^. Interestingly, we observed a significant association between *SCARB2* rs6812193 and PD in the Latino and indigenous Americas population, between *SCARB2* rs6825004 and PD in the Middle Eastern and Complex Admixture History populations. Besides this, we also identified one novel independent non-coding common genetic variant(rs11547135) in the European population. In the Ashkenazi Jewish population, rs530111925 at the *SCARB2* locus was identified as a novel, independent variant. Excessive genetic burden of *SCARB2* rare variants was not replicated in the European population. Previous studies had shown a possible association between *SCARB2* rare variants and PD risk with SKAT-O test^7,29^. This may be attributed to genetic and demographic differences among populations from different regions or lack of statistical power. Interestingly, *SCARB2* rare variants in exonic regions were found to be associated with PD risk in three other populations, the African American, the Ashkenazi Jewish, and the East Asian cohorts. Moreover, we found 5 novel single genetic variants (rs149997095, rs957776724, rs143518519, rs147159813 and rs35069772) significantly associated with increased PD risk in the African American population and none of which have previously been reported to be associated with PD in ClinVar. In the Ashkenazi Jewish cohort, single-variant-based tests showed that rs143655258 had a protective effect on PD. This variant results in an amino acid change, p. Met159Val, and has not been reported before in the context of PD risk. In the East Asian population, we identified two single significant missense variants, *SCARB2* rs773017713 and rs117600063, resulting in the p.Asp194Asn and p.Ile144Leu amino acid substitutions, which exhibit discordant effects on PD. Furthermore, rare variant analysis of *SCARB2* revealed mixed effect directions, with a balanced effect magnitude across the two directions. In such scenarios, the aggregate effect may appear null, despite the presence of biologically meaningful associations. The bidirectional effects have also been observed in genes like *TMEM175* and *CTSB*, where opposing variant effects can obscure the cumulative genetic burden on disease risk. These findings underscore a key limitation of conventional burden analyses that rely on the assumption of effect directionality. On the other hand, understanding such discordance was crucial as it suggested that targeting a single specific pathogenic variant—rather than the gene as a whole—may be a more effective strategy in clinical interventions.

### CTSB

We found *CTSB* rs1293298 as a significant risk factor only in the European and Complex Admixture History populations. Of note, *CTSB* rs1293298 in East Asians was excluded from the common variant analyses due to MAF <0.01. In the European cohort, we identified an independent common intronic variant (rs1293289), linked to the known GWAS hit *CTSB* rs1293298. In addition, *CTSB* rs73551266 was detected as a novel independent variant in the Latino and indigenous Americas population.

A recent study observed a nominal association between *CTSB* rare variants and PD risk in a European population^30^. However, our large European cohort (24,208 cases, 9,662 controls) did not replicate this result. In contrast to the previous study, we excluded carriers of *GBA1* variants from all cohorts to minimize potential bias, which may partly explain the discrepancy. Our findings further support the limitation of rare variant burden analysis, particularly in genes like *TMEM175* and *CTSB*, where both risk and protective variants may co-exist. Besides, future studies should focus on larger sample size in other populations to increase robustness.

### GBA1

We performed gene-based and single-variant-based tests in AMP-PD dataset to estimate genetic burden of rare variants in *GBA1* gene. Our results replicate and validate the established association with *GBA1*. We detected a genetic burden of *GBA1* rare variants on PD in the European population from the AMP-PD dataset, in concordance with results from previous work^7,31^. *GBA1* p. Asn409Ser was shown to be associated with PD risk on a single variant level. This specific mutation is classified as a “mild” mutation based on its phenotype in homozygous carriers with Gaucher disease^32,33^.

The associations of known GWAS hits with PD in other populations were observed although not reaching genome wide significance. This discrepancy may reflect distinct LD patterns, differences in haplotype structures among populations or limited statistical power due to smaller cohort size. Future haplotype-resolved analyses (e.g., phased whole-genome sequencing or ancestry-stratified fine-mapping) could clarify whether these associations arise from shared causal variants present on divergent haplotypes or population-specific functional variants embedded within risk haplotypes. On the other hand, our study illustrates heterogeneity in genetic associations across populations in PD. These combined analyses emphasize these three lysosomal related genes (*TMEM175, SCARB2* and *CTSB*) driving PD risk via both common variants and rare variants. Future research should prioritize detailed functional characterization of these novel variants and their potential patho-mechanism in diverse populations to uncover their roles, facilitating improved understanding and potential development of targeted interventions for PD.

The strength of this study is the analysis of a large-scale case-control cohort from the GP2 Neurobooster array with a population of European ancestry and 10 additional populations as well as WGS data from the AMP-PD dataset. There are some limitations. The observed replication may, in part, be influenced by partial sample overlap with the 2019 PD GWAS cohort. Notably, 7 out of the 126 cohorts included in our study were included in the previous Nalls’s PD GWAS. It is not possible to identify exactly which samples overlap, because the genotyping arrays are so different that kinship analysis does not identify related/duplicated samples. Despite this limitation, our study reinforces the association between lysosomal genes and PD risk in the European population. Additionally, the number of non-European individuals will need to be increased in future studies, being clearly smaller compared to the European cohort.

PD is a complex disorder with many genetic risk factors at multiple loci contributing to disease risk. Therefore, the novel variants reported in this study should be further investigated in terms of functional effects on PD pathogenesis in the future.

## Data Availability

Data used in the preparation of this article were obtained from the Global Parkinson's Genetics Program (GP2; https://gp2.org). Specifically, we used Tier 2 data from GP2 release 10 (https://doi.org/10.5281/zenodo.15748014). GP2 data are available on AMP PD (https://amp-pd.org).

https://amp-pd.org

## Author contributions

W.S. contributed to the design of the study, M.T. supervised the data analysis, W.S. contributed to detailed analytical workflows, data visualization and drafted the manuscript, C.S., M.T., T.G contributed to the guidance of the whole process and manuscript check.

## Acknowledgements

This project was supported by the Global Parkinson’s Genetics Program (GP2; https://gp2.org). GP2 is funded by the Aligning Science Across Parkinson’s (ASAP) initiative and implemented by The Michael J. Fox Foundation for Parkinson’s Research (MJFF). For a complete list of GP2 members see https://doi.org/10.5281/zenodo.7904831. For the purpose of Open Access, the author has applied a CC BY public copyright licence to any Author Accepted Manuscript version arising from this submission.

## Data and Code Availability Statement

Data used in the preparation of this article were obtained from the Global Parkinson’s Genetics Program (GP2; https://gp2.org). Specifically, we used Tier 2 data from GP2 release 10 (https://doi.org/10.5281/zenodo.15748014). GP2 data are available on AMP PD (https://amp-pd.org). All code generated for this article, and the identifiers for all software programs and packages used, are available on GitHub (https://github.com/GP2code/TMEM175_SCARB2_CTSB_PDrisk) and were given a persistent identifier via Zenodo (10.5281/zenodo.15799510).

