## Supplementary figures and tables for "*TMEM175, SCARB2* and *CTSB* associations with Parkinson’s disease risk across populations"

Supplementary tables and figures

Supplementary Table 1. Association analysis of *SCARB2* rs6812193 and *rs6825004* across all 11 populations

| *SCARB2* rs6812193(4:76277833:C: T) | | | | | *SCARB2* rs6825004(4:76189212:C: G) | | | |
| --- | --- | --- | --- | --- | --- | --- | --- | --- |
| Ancestry | MAF (cases) | MAF (controls) | P value | OR (95%CI) | MAF (cases) | MAF (controls) | P value | OR (95%CI) |
| AAC | 0.40 | 0.46 | 0.43 | 0.86(0.72-1.03) | 0.23 | 0.21 | 0.96 | 1.01(0.78-1.30) |
| AFR | 0.46 | 0.45 | 0.76 | 1.02(0.90-1.15) | 0.18 | 0.19 | 0.45 | 1.06(0.91-1.25) |
| AJ | 0.29 | 0.33 | 0.075 | 0.82(0.65-1.02) | 0.40 | 0.39 | 0.77 | 1.03(0.84-1.27) |
| AMR | 0.22 | 0.15 | 0.020* | 0.83(0.71-0.97) | 0.49 | 0.43 | 0.69 | 1.02(0.91-1.15) |
| CAS | 0.21 | 0.21 | 0.69 | 1.05(0.83-1.32) | 0.32 | 0.37 | 0.12 | 0.85(0.69-1.04) |
| EAS | 0.076 | 0.078 | 0.27 | 0.90(0.74-1.09) | 0.36 | 0.34 | 0.23 | 1.07(0.96-1.19) |
| EUR | 0.35 | 0.36 | 7.81e-03* | 0.94(0.90-0.98) | 0.30 | 0.31 | 0.036* | 0.95(0.91-1.00) |
| FIN | 0.33 | 0.27 | 0.43 | 2.51(0.25-24.81) | 0.28 | 0.36 | 0.38 | 0.41(0.055-3.00) |
| MDE | 0.42 | 0.41 | 0.48 | 0.94(0.78-1.12) | 0.37 | 0.39 | 0.023* | 0.80(0.66-0.97) |
| SAS | 0.27 | 0.29 | 0.39 | 0.85(0.60-1.22) | 0.23 | 0.23 | 0.92 | 0.98(0.69-1.39) |
| CAH | 0.31 | 0.35 | 0.15 | 0.84(0.67-1.07) | 0.29 | 0.28 | 0.042* | 0.78(0.62-0.99) |

Abbreviations: AAC, African American; AFR, African; AJ, Ashkenazi Jewish; AMR, Latino and indigenous Americas; CAS, Central Asian; EAS, East Asian; EUR, European; FIN, Finnish population isolate; MDE, Middle Eastern; SAS, South Asian; CAH, Complex Admixture History. MAF, minor allele frequency. OR, odds ratio. CI, Confidence Interval. NA, not available. * means p value < 0.05.

Location of variants are based on GRCh38/hg38.

Supplementary Table 2. Association analysis of *CTSB* rs1293298 across all 11 populations

| Ancestry | MAF (cases) | MAF (controls) | P value | OR (95%CI) |
| --- | --- | --- | --- | --- |
| AAC | 0.29 | 0.33 | 0.20 | 0.86(0.68-1.08) |
| AFR | 0.33 | 0.31 | 0.26 | 1.08(0.95-1.23) |
| AJ | 0.29 | 0.31 | 0.29 | 0.88(0.71-1.11) |
| AMR | 0.14 | 0.081 | 0.69 | 0.96(0.79-1.16) |
| CAS | 0.11 | 0.13 | 0.46 | 0.89(0.66-1.21) |
| EAS | NA | NA | NA | NA |
| EUR | 0.24 | 0.25 | 5.82e-04* | 0.92(0.87-0.96) |
| FIN | 0.21 | 0.23 | 0.31 | 0.29(0.026-3.21) |
| MDE | 0.27 | 0.28 | 0.53 | 0.94(0.76-1.15) |
| SAS | 0.17 | 0.17 | 0.94 | 1.02(0.67-1.55) |
| CAH | 0.21 | 0.26 | 0.021* | 0.73(0.56-0.95) |

Abbreviations: AAC, African American; AFR, African; AJ, Ashkenazi Jewish; AMR, Latino and indigenous Americas; CAS, Central Asian; EAS, East Asian; EUR, European; FIN, Finnish population isolate; MDE, Middle Eastern; SAS, South Asian; CAH, Complex Admixture History. MAF, minor allele frequency. OR, odds ratio. CI, Confidence Interval. NA, not available. * means p value < 0.05.

Location of variants are based on GRCh38/hg38.

Supplementary Table 3. Burden test and Skat test *SCARB2* with GP2 dataset based on significant results from Skat-O analysis.

| Ancestry | Variants | Test | NumVar | P value |
| --- | --- | --- | --- | --- |
| AAC | exonic | SKAT | 21 | 8.90e-04* |
| AJ | exonic | SKAT | 5 | 0.021* |
| EAS | exonic | SKAT | 17 | 0.013* |
|  | Nonsyn and LoF | SKAT | 12 | 0.011* |

Abbreviations: EUR, European; EAS, East Asian; SAS, South Asian; CAS, Central Asian; AMR, Latino and indigenous Americas; Nonsyn, non-synonymous variants; LoF, and Loss of function; NumVar, number of variants.

Supplementary Table 4A. Skat-O analysis of *GBA1* with AMP-PD

|  | variants | AMP-PD | | |
| --- | --- | --- | --- | --- |
| Ancestry |  | NumVar | rho | P value |
| EUR | exonic | 41 | 1 | 8.60e-03* |
|  | Nonsyn and LoF | 28 | 1 | 0.020* |

Abbreviations: EUR, European; NumVar, number of variants.

Supplementary Table 4B. Burden test of *GBA1* with AMP-PD dataset based on significant results from Skat-O analysis.

| Gene | Ancestry | Variants | Test | NumVar | P value | Beta |
| --- | --- | --- | --- | --- | --- | --- |
| *GBA1* | EUR | exonic | Fp | 42 | 1.40e-03* | / |
|  |  |  | CMCwald |  | 7.30e-03* | 0.42 |
|  |  |  | MadsonBrowning |  | 4.00e-04* | / |
|  |  |  | Zeggini |  | 0.011* | / |
|  |  |  | CMC |  | 7.10e-03* | / |
|  |  | Nonsyn and LoF | Fp | 28 | 4.70e-03* | / |
|  |  |  | CMCwald |  | 0.012* | 0.42 |
|  |  |  | MadsonBrowning |  | 4.60e-03* | / |
|  |  |  | Zeggini |  | 0.012* | / |
|  |  |  | CMC |  | 0.012* | / |

Abbreviations: EUR, European; Nonsyn, non-synonymous variants; LoF, and Loss of function; NumVar, number of variants.

Supplementary Table 4C. Significant signal of *GBA1* with single variant burden analysis

| Gene | Dataset | Ancestry | Variant | Category | MAF | Direction | Effect | SE | P value |
| --- | --- | --- | --- | --- | --- | --- | --- | --- | --- |
| *GBA1* | AMP-PD | EUR | rs76763715  (1:155235843) | missense | 3.24e-03 | + | 1.11 | 0.39 | 4.00e-03* |

Abbreviations: EUR, European; MAF, minor allele frequency; SE, standard error. + means risk, - means protective.

Supplementary Table 5. Skat-O analysis of *TMEM175* with p.M393T and p.Q65P included as covariates.

| *TMEM175* | | | | |
| --- | --- | --- | --- | --- |
| Ancestry | Variants | Num Var | rho | P value |
| AAC | exonic | 59 | 0 | 0.62 |
|  | Nonsyn and LoF | 38 | 0 | 0.62 |
| AFR | exonic | 76 | 1 | 0.87 |
|  | Nonsyn and LoF | 47 | 1 | 0.87 |
| AJ | exonic | 24 | 0 | 0.24 |
|  | Nonsyn and LoF | 15 | 0 | 0.24 |
| AMR | exonic | 72 | 0 | 0.53 |
|  | Nonsyn and LoF | 42 | 1 | 0.29 |
| CAS | exonic | 39 | 1 | 0.69 |
|  | Nonsyn and LoF | 26 | NA | NA |
| EAS | exonic | 52 | 1 | 0.26 |
|  | Nonsyn and LoF | 34 | 1 | 0.089 |
| EUR | exonic | 128 | 0 | 0.12 |
|  | Nonsyn and LoF | 69 | 0 | 0.14 |
| FIN | exonic | 4 | NA | NA |
|  | Nonsyn and LoF | 2 | NA | NA |
| MDE | exonic | 47 | 0 | 0.61 |
|  | Nonsyn and LoF | 24 | 0 | 0.61 |
| SAS | exonic | 25 | 0 | 0.61 |
|  | Nonsyn and LoF | 18 | 1 | 0.61 |
| CAH | exonic | 47 | 1 | 0.27 |
|  | Nonsyn and LoF | 30 | 1 | 0.012* |

Abbreviations: AAC, African American; AFR, African; AJ, Ashkenazi Jewish; AMR, Latino and indigenous Americas; CAS, Central Asian; EAS, East Asian; EUR, European; FIN, Finnish population isolate; MDE, Middle Eastern; SAS, South Asian; CAH, Complex Admixture History; Nonsyn, non-synonymous variants; LoF, and Loss of function; NA, not available. NumVar, number of variants.

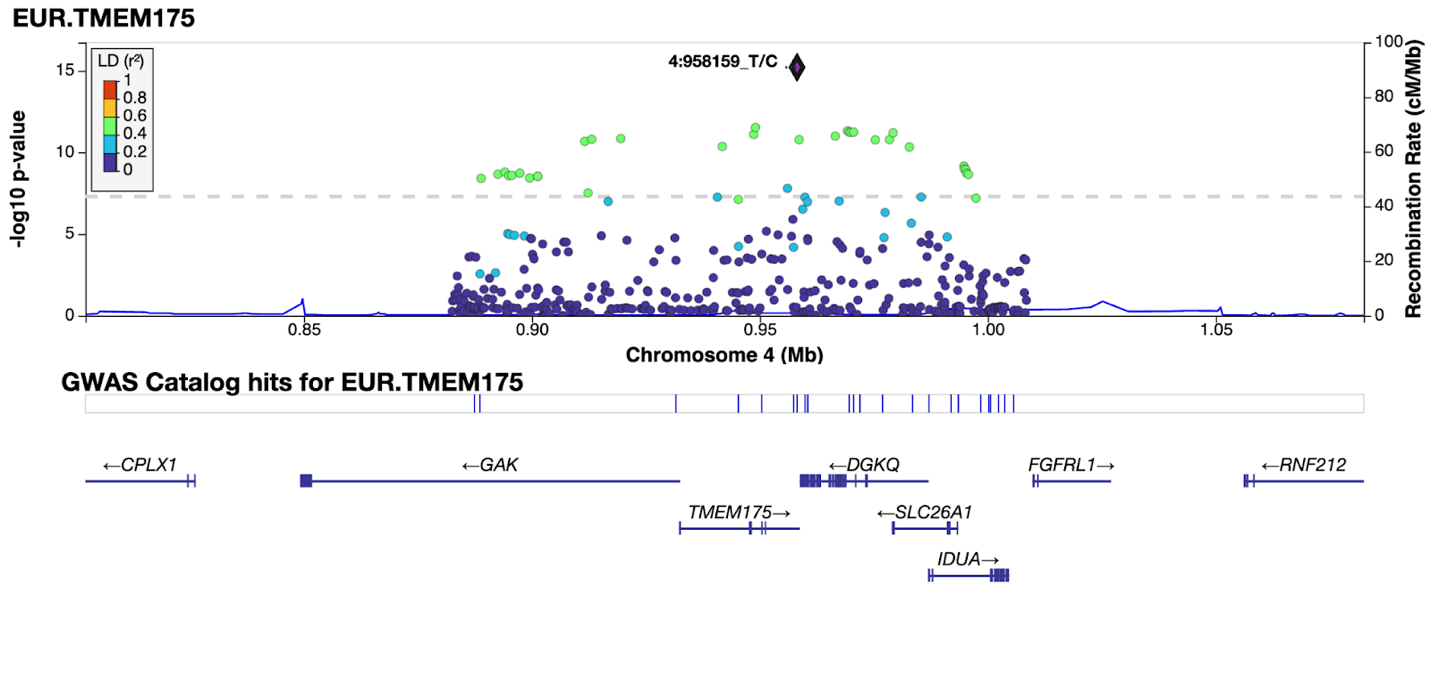

Figure 1: The most associated signal, rs34311866(4:958159), in *TMEM175* locus in GP2-EUR cohort.

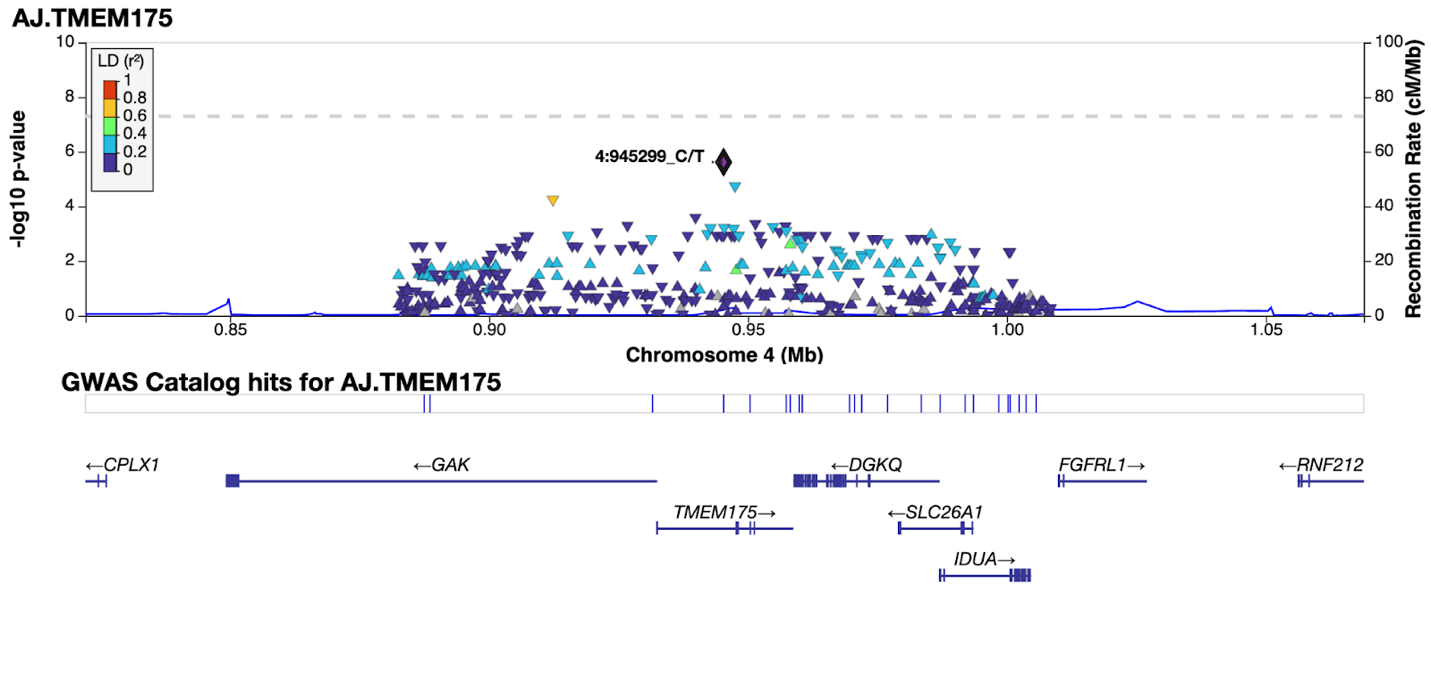

 Figure 2: *TMEM175* association with Parkinson’s disease in GP2-AJ cohort: the most associated signal, rs6599388 (4:945299).

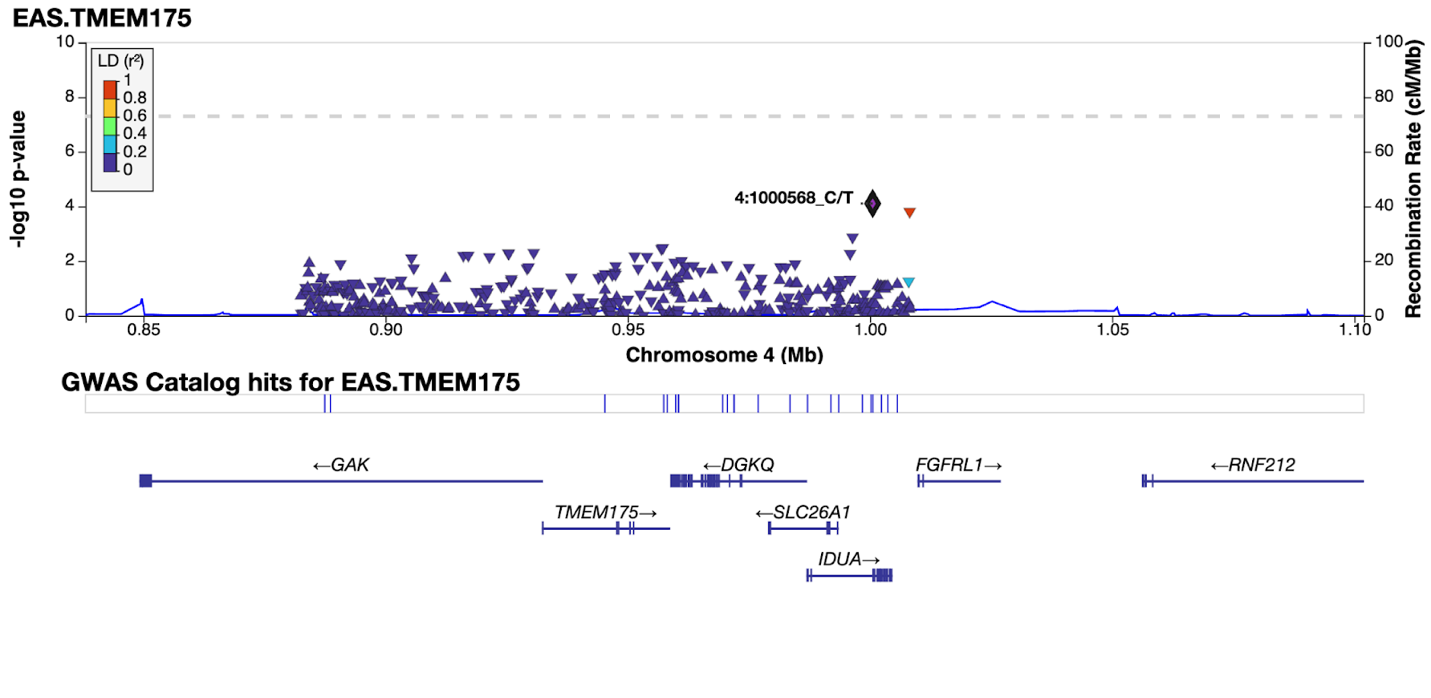

Figure 3: *TMEM175* association with Parkinson’s disease in GP2-EAS cohort: the most associated signal, rs3755956 (4:1000568).

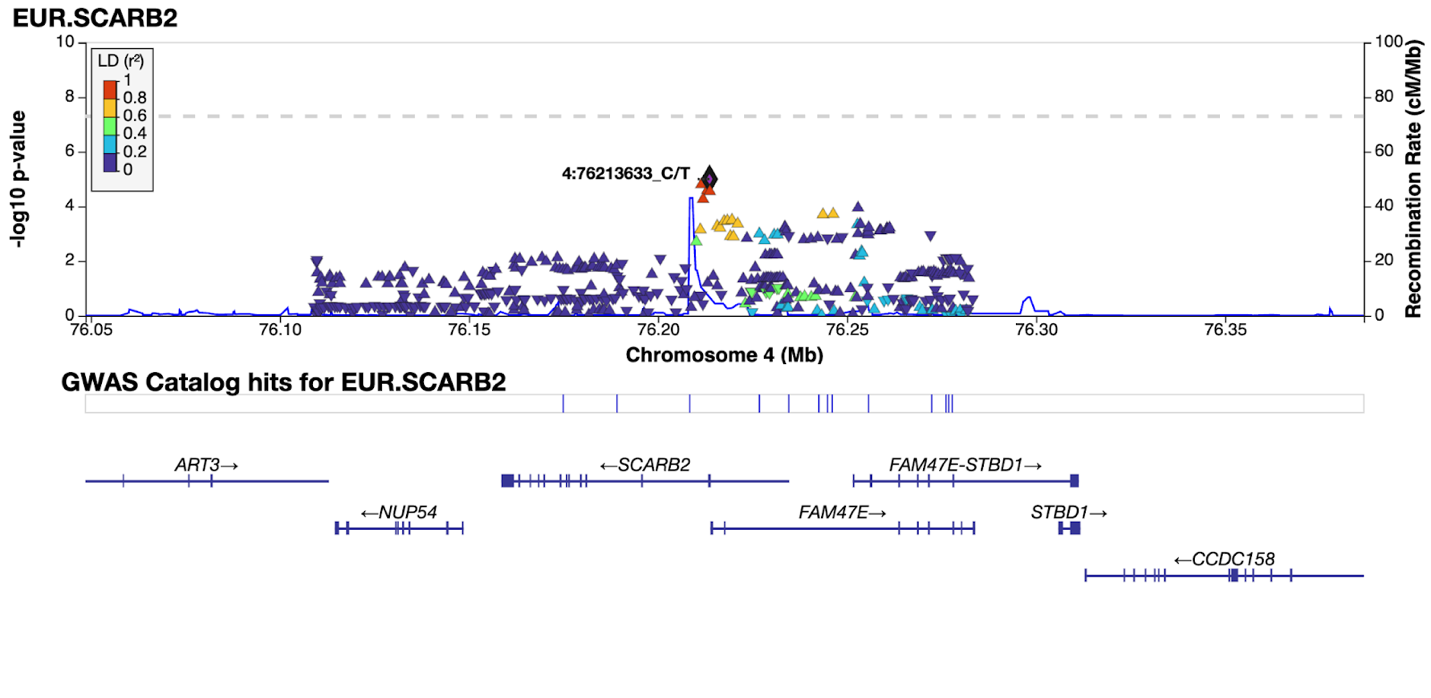
Figure 4: The most associated signal, [rs11547135](https://www.ncbi.nlm.nih.gov/snp/rs11547135)(4:76213633), within SCARB2 locus in GP2-EUR cohort.

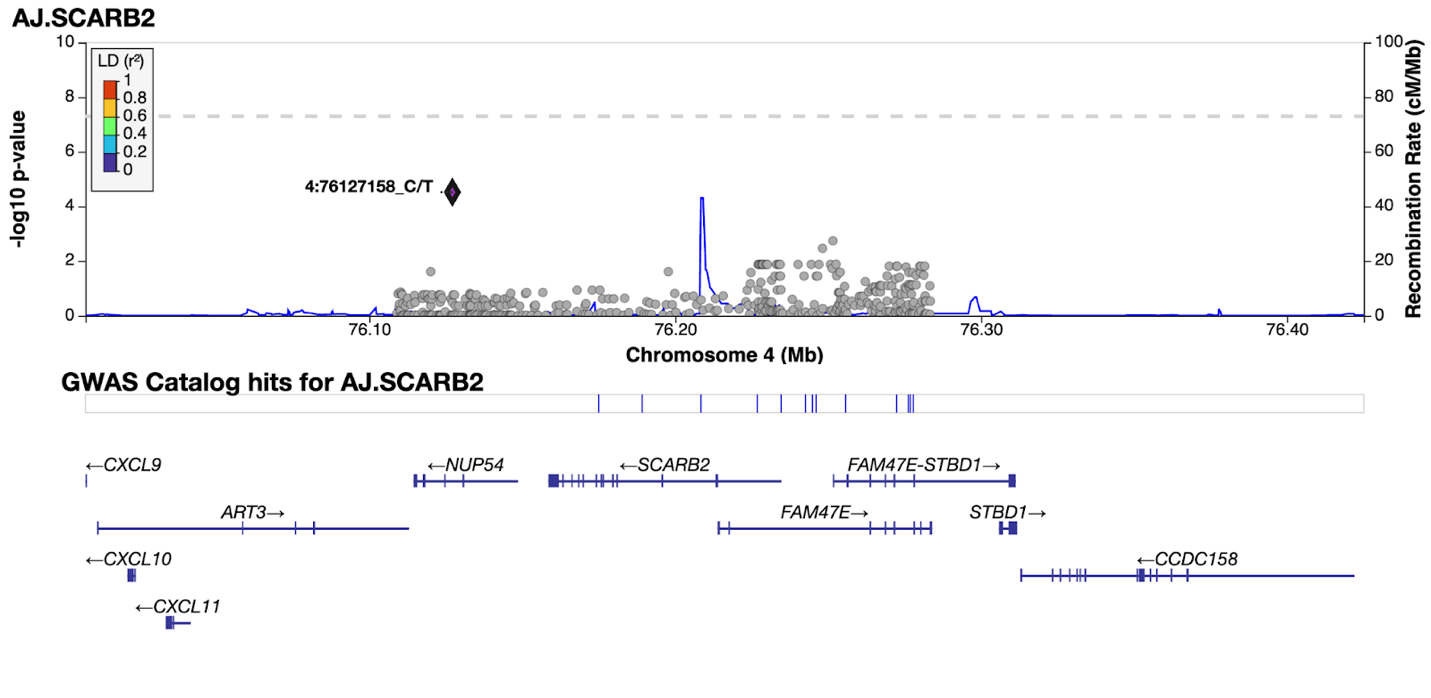

Figure 5: The most associated signal, [rs530111925](https://www.ncbi.nlm.nih.gov/snp/rs530111925)(4:76127158), within SCARB2 locus in GP2-AJ cohort.

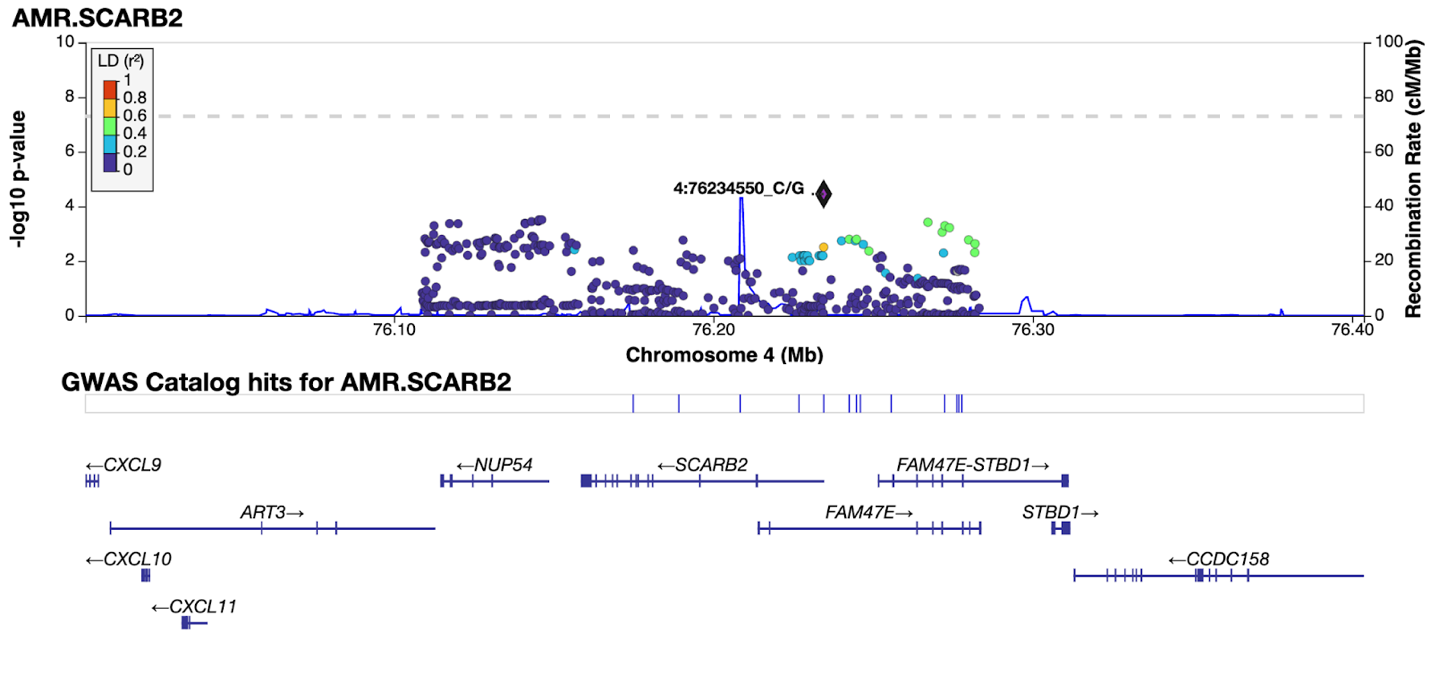

Figure 6: The most associated signal, [rs73828719](https://www.ncbi.nlm.nih.gov/snp/rs73828719)(4:76234550), within SCARB2 locus in GP2-AMR cohort.

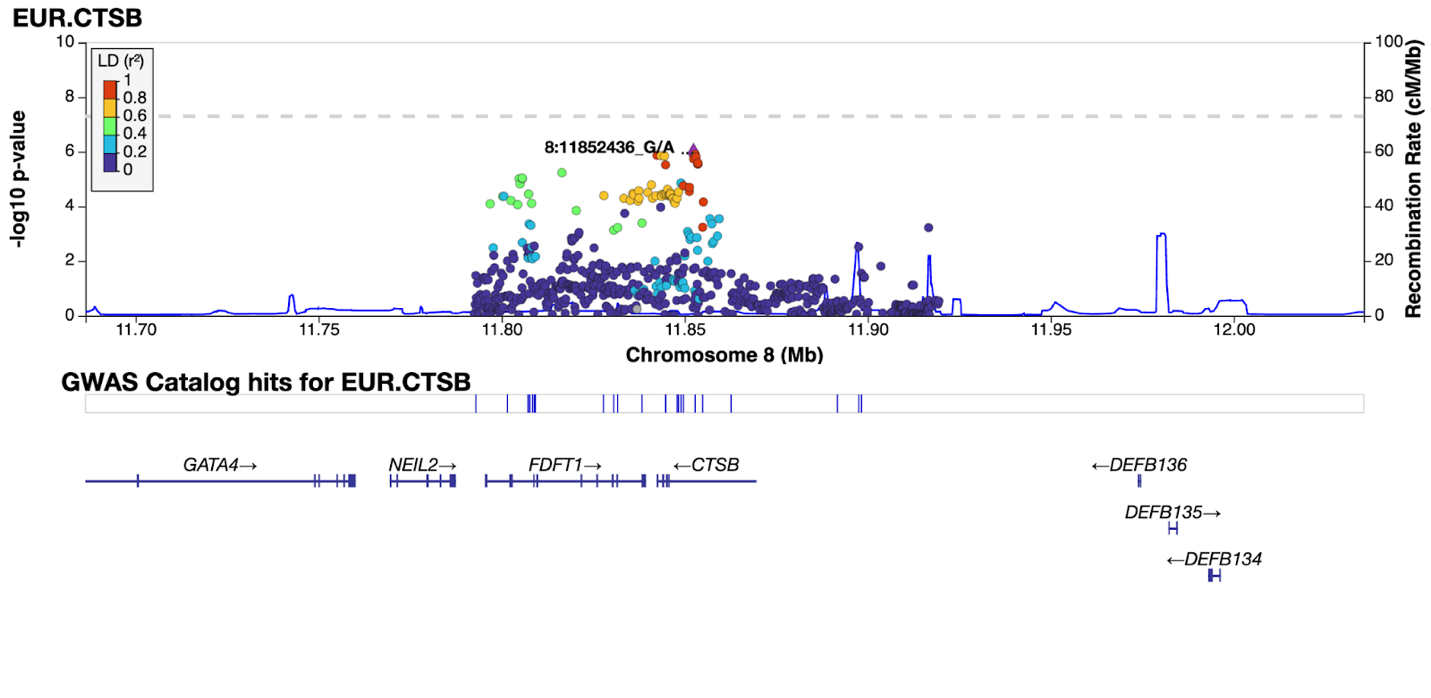

Figure 7: *CTSB* association with Parkinson’s disease:the most associated signal, rs1293289(8:11852436) in GP2-EUR cohort.

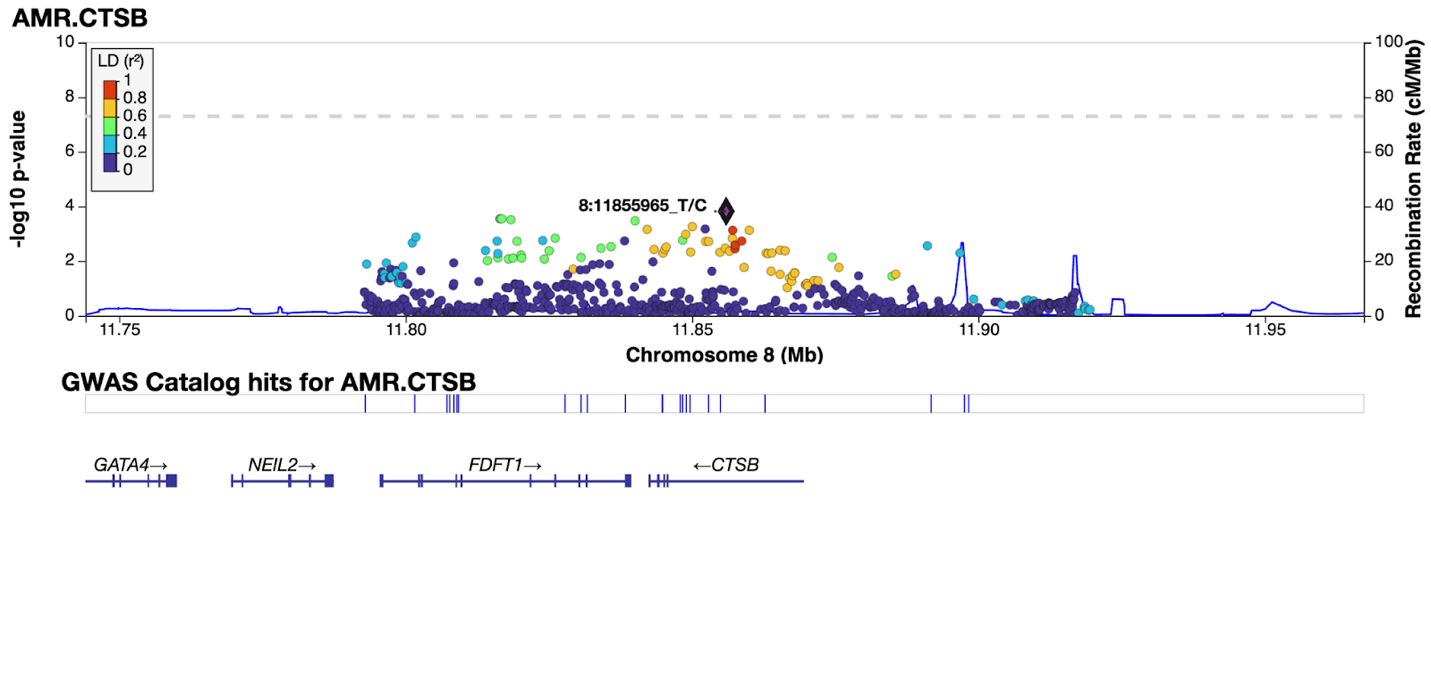

Figure 8: *CTSB* association with Parkinson’s disease: the most associated signal, rs73551266 (8:11855965) in GP2-AMR cohort.

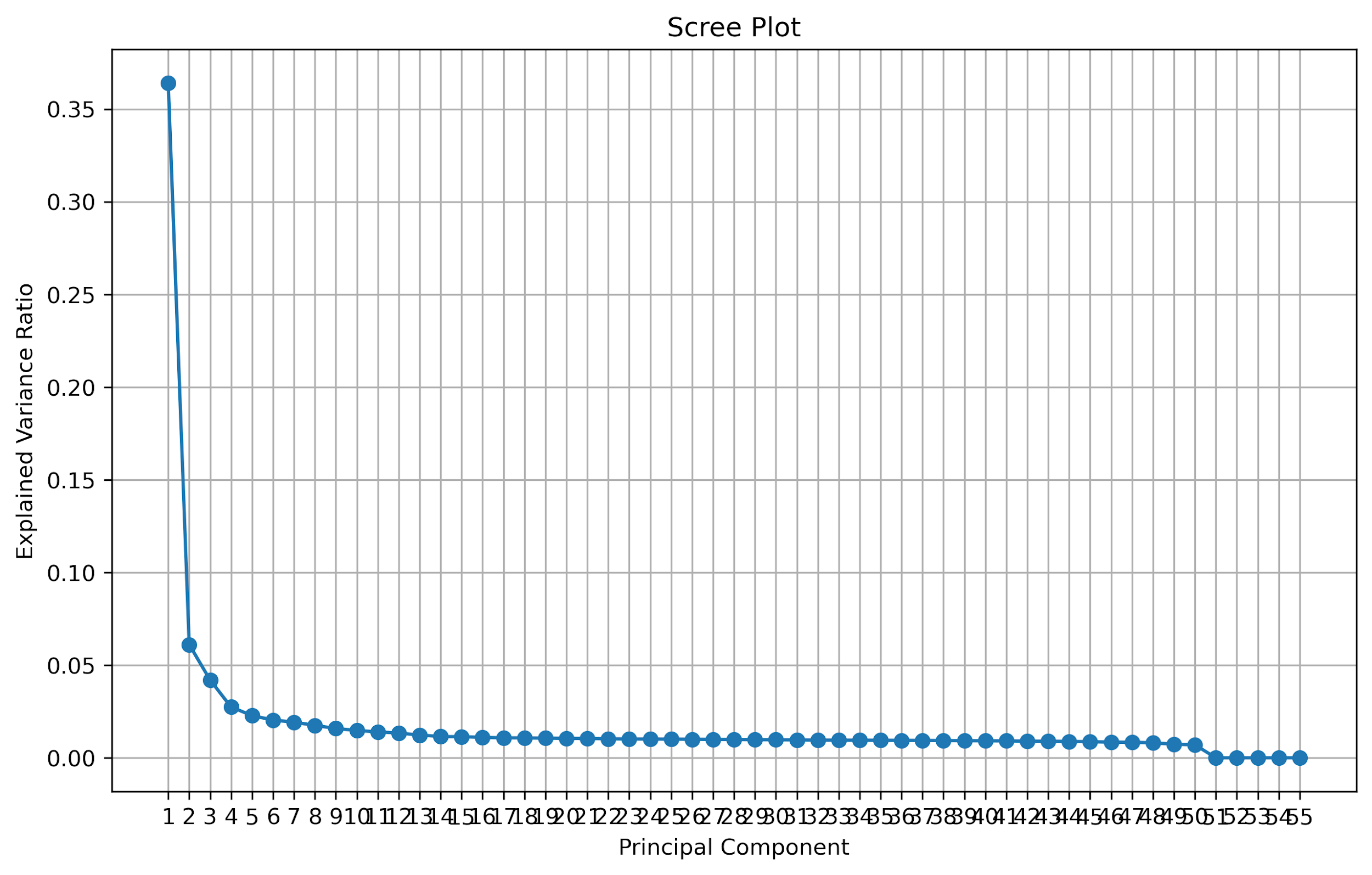

Figure 9: Scree plot of principle components in European population.
